# Reporting Quality of Mendelian Randomization Studies Based on STROBE-MR: An Investigation Powered by Large Language Models

**DOI:** 10.64898/2026.10.21.26366143

**Authors:** Jiangjie Zhou, Xuyutian Wang, Zhaozhen Xu, Edwin Simpson, Julian PT Higgins, Tom R Gaunt, Yi Liu

## Abstract

2

**Background:** Mendelian randomization (MR) has been widely used to investigate causal relationships between exposures and outcomes. However, concerns have also been raised about the reporting quality of MR studies. STROBE-MR was developed as a reporting guideline for MR studies, but assessments of compliance with STROBE-MR in published MR studies remain limited and rely largely on manual review. With the rapid development of large language models (LLMs), it is now possible to automate STROBE-MR compliance assessment.

**Methods:** We developed a pipeline using OpenAI’s GPT-5-mini model for automated assessment of MR reporting quality. Model performance was evaluated on an annotated dataset of 46 eligible MR articles. We applied the pipeline to more than 13,000 MR articles published between 2005 and 2025 to assess compliance with the STROBE-MR guideline and to explore reporting patterns across STROBE-MR items, sections, and time periods. Complete compliance required reporting all applicable components of an item; a sensitivity analysis used a partial compliance criterion requiring at least one applicable component.

**Results:** Our LLM-based pipeline agreed with the human consensus on 89.0% of the STROBE-MR assessments. On average, 22 of 43 items (51.2%) met the complete compliance criterion, with substantial variation in reporting rates across STROBE-MR items and sections. The least reported items were found in the “Methods”, “Results”, and “Other Information” sections. The number of reported items remained stable over time, with slight decreases after 2021. Sensitivity analysis showed that temporal trend patterns differed between complete and partial compliance, particularly in the Methods, Discussion and Other Information sections.

**Conclusions:** This study demonstrates the potential of LLMs to automate reporting quality assessment in MR studies. Our results suggest that reporting quality still needs to be improved, and further efforts are required to promote the use of STROBE-MR. Our findings also suggest potential refinements to the STROBE-MR guideline to support more consistent application.

**Key Points:** *Question:* Can large language models (LLMs) accurately assess adherence to the STROBE-MR reporting guideline in Mendelian randomization (MR) studies, and what patterns can we observe in MR reporting?

*Findings:* The LLM-based approach achieved good agreement with human consensus (89.0%) and identified substantial variation in reporting quality across STROBE-MR items and sections. No improvement in number of items reported was observed since the introduction of the STROBE-MR guideline.

*Meaning:* LLMs can enable scalable assessment of reporting guideline compliance, providing a practical approach for monitoring reporting quality. Further efforts are needed to promote the use of reporting guidelines for MR studies.

## 3 Introduction

Mendelian randomization (MR) is an analytical method that uses genetic variants, such as single-nucleotide polymorphisms (SNPs), as instrumental variables (IVs) to infer causal relationships between exposures and outcomes, thereby reducing unmeasured confounding that often biases traditional observational studies [1, 2, 3]. Over the past two decades, extensions of MR methods [4, 5, 6], software packages [7], and public datasets [8] have emerged. The number of published MR studies has also increased substantially [9], reflecting the growing popularity of the approach in epidemiology and biomedical research. However, concerns have been raised about the completeness, transparency, and reproducibility of MR reporting [10]. Many studies provide insufficient methodological detail, which limits their interpretability and undermines the credibility of causal claims [11, 12]. For example, IVs used in an MR study must satisfy three core assumptions: relevance, independence, and exclusion [13]. Authors should therefore report whether these assumptions were assessed and whether the results were robust to violations of the assumptions. As two-sample MR studies have become more common, additional concerns have also been raised about the comparability of genetic variant-exposure associations between the exposure and outcome samples. To address these issues, the STROBE-MR (Strengthening the Reporting of Observational Studies in Epidemiology for Mendelian Randomization) guideline was developed[10]. The guideline contains 20 main items containing 43 sub-items across six sections: Title and Abstract, Introduction, Methods, Results, Discussion, and Other Information. We work mainly with the 43 sub-items in this paper and hereafter refer to them as ‘items’ for simplicity.

STROBE-MR provides a structured framework to help authors report MR studies clearly. It also offers reviewers and editors a useful tool for assessing and identifying studies with high-quality reporting[14, 15]. Despite the guideline’s availability, assessments of MR reporting quality remain largely manual, relying on expert reviewers to evaluate adherence to STROBE-MR criteria [16]. For example, one study [17] assessed MR reporting manually and concluded that the quality of MR articles needs to be improved, but its work was limited to 116 articles using UK Biobank data. This manual approach limits large-scale assessment of MR reporting. Consequently, there is a need for automated methods that can efficiently and accurately evaluate reporting quality at scale.

Recent advances in large language models (LLMs) offer new opportunities: these models can process and interpret complex biomedical texts efficiently, demonstrating strong potential in biomedical knowledge extraction [18], literature selection [19], risk of bias assessment [20, 21], and reporting quality assessment [22, 23].

In this study, we developed an LLM-based approach to address the challenges of large-scale reporting quality assessment and evaluated its performance on an annotated dataset. We collected full texts and supplementary materials for more than 13,000 MR studies published between 2007 and 2025 and applied this approach to analyze reporting trends across different STROBE-MR items, sections, and time periods. Unlike previous studies that classified complex STROBE-MR items as “partially reported” [17], we decomposed compound items into more granular components, enabling a more detailed and informative assessment of reporting quality.

This project aims to leverage LLMs to develop and validate an automated system capable of rapidly assessing the reporting quality of MR publications based on the STROBE-MR guideline. Such a system would enable large-scale evaluation of reporting practices across the MR literature and help evidence synthesis researchers screen MR articles with transparent reporting. It also has the potential to help authors improve the completeness and reproducibility of their research papers and to support journals and editors in identifying reporting problems early in the publication process.

## 4 Methods

### 4.1 Data collection

We used PubMed Central (PMC) as our main data source. First, a database query was performed to retrieve PMCIDs for Mendelian randomization studies. We used the following query for this purpose:

“Mendelian Randomization Analysis”[Mesh] OR “Mendelian Randomi*ation”[Title/Abstract]

A total of 13,568 results were returned (2025-11-5). A Python script (*data_collection*.*py*) was developed to download the corresponding HTML files (including full texts of articles) and supplementary materials, and a second Python script was developed to extract information and format each article into a JSON object. Fields included in the JSON object are listed in Table S1-3. All JSON objects were stored in a MongoDB database.

Supplementary materials can have different file types, and support for non-text data by non-multimodal LLMs may be inadequate. To provide appropriate context to the model, we used the following rules to extract text from them (*text_extract*.*ipynb*):

1. If it is a figure (png, jpeg, svg, etc.): Extract its caption and description if available
2. If it is a document (pdf, doc, docx, txt, etc.): Extract all the text included in the document (tables were treated as plain text). If figures are included in the document, follow rule 1.
3. If it is a data set (csv, tsv, xls, xlsx): Extract the first 10 rows of each table to limit the length of the LLM prompt. If the figures are included, follow rule 1.

### 4.2 STROBE-MR item decomposition

Some STROBE-MR items are compound questions that contain requirements for different types of information, for example, item 4(a): *Setting: Describe the study design and the underlying population, if possible. Describe the setting, locations, and relevant dates, including periods of recruitment, exposure, follow-up, and data collection, when available*. A MR article can sometimes only present part of this information, so some information can be lost if we only give a simple “yes” or “no” answer for this item. To avoid losing this information, a previous study[17] allowed coding of this item as “partially” reported.

In this study, to make our assessments more detailed, we divided compound items into more granular components that can be assessed independently. For example, we can divide item 4(a) into components of study design, population, locations etc. We defined 80 components from the original 43 STROBE-MR items. The definitions of these components can be found in Table S1-1. A mapping process was used to map components back to items, as described in the Evaluation section below.

### 4.3 Prompt Design and Assessment

We used OpenAI’s GPT models (including GPT-5 and GPT-5-mini) for the assessment. For each MR article, the main body of text, text extracted from supplementary materials, and the STROBE-MR checklist were fed into the LLM as inputs (text, supp and strobe respectively in the example in Table 1). We then prompted the LLM to determine the type of the article. Since STROBE-MR applies only to original research, we prompted the LLM to ignore the article if it was not an original research article (for example, a literature review). For original research articles, the LLM should decide whether the article described a one-sample or two-sample design. The LLM should then assess whether it has reported each STROBE-MR component and give reasons for that decision. The assessment output was converted into a list of JSON objects. Each JSON object contained 4 elements:

- index: the STROBE-MR component being assessed (numbered 1 to 80)
- status: 1 if the article reported information required by the component, 0 if it does not, 2 if this component is not applicable to the article (for example, components for two-sample MR are not applicable to one-sample designs)
- rationale: step-by-step reasoning to explain why the LLM assigned the status
- relevant_text: relevant text from the article to support the decision.

**Table 1:** LLM prompt template for OpenAI platform.

| Role | Prompt |
| --- | --- |
| System | You are a highly skilled medical research assistant with extensive knowledge of mendelian randomization and STROBE guidelines. Your task is to assess whether specific mendelian randomization articles meet the given STROBE criteria. For each criterion, you will analyze the article text and provide a detailed justification for your assessment. |
| User | # Task: Your job is to assess whether the given article meets the specified STROBE criterion and provide justification for your assessment.<br># Article: Below is the article text:<br>{text}<br># Supplementary Materials:<br>{supp}<br># Before proceeding to the assessment, decide whether the article is an original research article. If it is not (for example, a review or comment), respond with an empty list: []<br># STROBE-MR Criterion:<br>Format (CSV):<br>index: int-The index of STROBE-MR item being assessed.<br>section: str-The section of STROBE-MR item being assessed.<br>item: str-The specific STROBE-MR item being assessed.<br>note: str-Additional notes regarding the STROBE-MR item.<br>Whole STROBE-MR Checklist:<br>{strobe}<br># Before start:<br>Decide publication type of this article (i.e., original research, review, comment, etc.). If it is not an original research article, skip this article and leave other fields with empty list or string.<br>If it is an original research article, decide its study design (e.g., two-sample MR, one-sample MR, multivariable MR, network MR, non-linear MR, etc.) and use this information to inform your assessment of the STROBE-MR criteria.<br># Assessment: Given the criterion items above, use the article text and supplementary materials to determine whether the article meets these criteria.<br>Think step by step and justify your answer. Format your response as a JSON objects, Example JSON output:<br>{<br>pub_type: original research,<br>study_design: two-sample MR,<br>assessments: [{<br>index: int-The index of STROBE-MR item being assessed.<br>rationale: str-Your reasoning as to why the article does or does not meet that criterion.<br>relevant_text: str-The specific text from the article that is relevant to assessing this criterion, empty string if no relevant text is found or not applicable.<br>status: int-1 if the article meets criterion, 0 if it does not, 2 if this item is not applicable to the article.<br>}]<br>} |

We used OpenAI’s Responses API^1^ to generate structured JSON output (JSON schema using pydantic and other parameters are listed in Table S1-2). A complete prompt can be found in Table 1 and settings.py. The implementation of this assessment process can be found at *mr_exp_large*.*ipynb*.

### 4.4 Evaluation

We randomly sampled 50 articles as the evaluation set. Four articles were excluded because MR was not the main purpose or they did not have human participants, leaving 46 articles for evaluation (four studies used a one-sample design, while the remaining studies used a two-sample design). Two reviewers (JZ, XW) independently assessed their compliance with the STROBE-MR checklist (A tool named *AnnotationToolMR* was developed to assist this task). For each STROBE-MR component, the reviewers answered with either “reported”, “not reported”, or “not applicable”. If two reviewers disagreed with each other on one component, all authors discussed and resolved the disagreement to establish a consensus rating for this component. Model-human consensus agreement was the proportion of model ratings that matched these consensus ratings. We then compared the model-human consensus agreement of GPT-5 and GPT-5-mini on the evaluation set and we selected the smaller model for the final assessment of the whole data set if there was no significant performance difference.

To summarize the STROBE-MR compliance across the whole data set, we calculated the average percentage of articles that met STROBE-MR by section, item, and component, respectively. Components that were not applicable to the article were ignored in these calculations. In the primary analysis, the 80 smaller components were mapped to the 43 STROBE-MR items by defining complete compliance as reporting all applicable components of an item. In practice, authors of MR research could have different interpretations of the STROBE-MR items, and may believe that some components are not necessary. We therefore conducted a sensitivity analysis using a partial compliance criterion, defined as reporting at least one applicable component of an item, to explore how an alternative mapping strategy could influence the results. This criterion includes items with complete compliance. Analysis code can be found at *analysis_exp_large*.*ipynb*

## 5 Results

The full texts of 13,434 articles published from 2005 to 2025 were collected from PMC. 10,919 of these included supplementary files. After evaluation by the LLM, 12,699 articles were found to be original MR research articles and passed to our STROBE-MR compliance assessment pipeline. Most of these (88.6%) used two-sample designs. 10,641 studies were published after 2021 (when the STROBE-MR was first introduced) and 2,789(26.2%) of these mentioned or cited STROBE-MR.

### 5.1 Model Evaluation

Of the 46 studies included in the final evaluation set, 4 used one-sample designs, whereas the remaining 42 used two-sample designs. 36 studies were published after 2021, and 10 of these (27.8%) explicitly claimed compliance with STROBE-MR. However, no differences in the average number of items reported were found between studies with and without self-claimed compliance (both groups reported an average of 24 items). Table 2 shows the distribution of grades in each STROBE-MR section by the two reviewers. Reviewers have a maximal percentage of disagreement of 10.2% and most of the disagreements occurred in the Methods and Results sections. Detailed results for each STROBE-MR component can be found in Table S1-4.

**Table 2:** Annotation Summary by STROBE-MR Section.

| Section | JZ |  |  | XW |  |  | Consensus |  |  | Disagreement | Total |
| --- | --- | --- | --- | --- | --- | --- | --- | --- | --- | --- | --- |
|  | Reported | NR | NA | Reported | NR | NA | Reported | NR | NA |  |  |
| Title and Abstract | 46 (100.0%) | 0 (0.0%) | 0 (0.0%) | 46 (100.0%) | 0 (0.0%) | 0 (0.0%) | 46 (100.0%) | 0 (0.0%) | 0 (0.0%) | 0 (0.0%) | 46 |
| Introduction | 88 (95.7%) | 4 (4.3%) | 0 (0.0%) | 89 (96.7%) | 3 (3.3%) | 0 (0.0%) | 88 (95.7%) | 4 (4.3%) | 0 (0.0%) | 1 (1.1%) | 92 |
| Methods | 1067 (58.0%) | 756 (41.1%) | 17 (0.9%) | 1097 (59.6%) | 719 (39.1%) | 24 (1.3%) | 1152 (62.6%) | 670 (36.4%) | 18 (1.0%) | 183 (9.9%) | 1840 |
| Results | 504 (57.7%) | 337 (38.6%) | 33 (3.8%) | 515 (58.9%) | 338 (38.7%) | 21 (2.4%) | 524 (60.0%) | 324 (37.1%) | 26 (3.0%) | 89 (10.2%) | 874 |
| Discussion | 406 (73.6%) | 146 (26.4%) | 0 (0.0%) | 424 (76.8%) | 128 (23.2%) | 0 (0.0%) | 423 (76.6%) | 129 (23.4%) | 0 (0.0%) | 20 (3.6%) | 552 |
| Other Information | 135 (48.9%) | 139 (50.4%) | 2 (0.7%) | 148 (53.6%) | 121 (43.8%) | 7 (2.5%) | 138 (50.0%) | 138 (50.0%) | 0 (0.0%) | 27 (9.8%) | 276 |

NR: not reported, NA: not applicable.

GPT-5 and GPT-5-mini agreed with the human consensus on 87.3% and 89.0% of component assessments, respectively. As GPT-5-mini showed a slightly higher performance (and has a lower cost) we selected this model for the assessment of whole dataset. Although the overall accuracy was high, it could decrease for items in the Methods and Results sections (see Table S1-5).

### 5.2 General Results

On average, 22 of the 43 items of STROBE-MR met the complete compliance criterion. The most frequently reported items were item 14: key results (99.3%), item 1: *title and abstract* (98.5%), item 16a: *meaning of results* (97.9%), item 2: *background* (97.3%), item 16c: *clinical relevance* (97.3%), and item 8: *sensitivity analysis* (96.9%). However, item 11c: *translating to absolute risk* (2.2%), item 9b: *registration of study* (2.4%), item 4b: *participants* (5.6%), item 18: *funding* (6.3%), and item 6c: *details of MR estimator* (8.7%) were rarely reported among MR articles. Details can be found in Figure 1, results for each STROBE-MR component can be found in Table S1-6. When comparing number of items reported across years, we found it remained stable around 24 items. (Figure S2-1). However, the mean reporting number decreased from 24 items in 2021 to 21 items in 2025.

**Figure 1:**
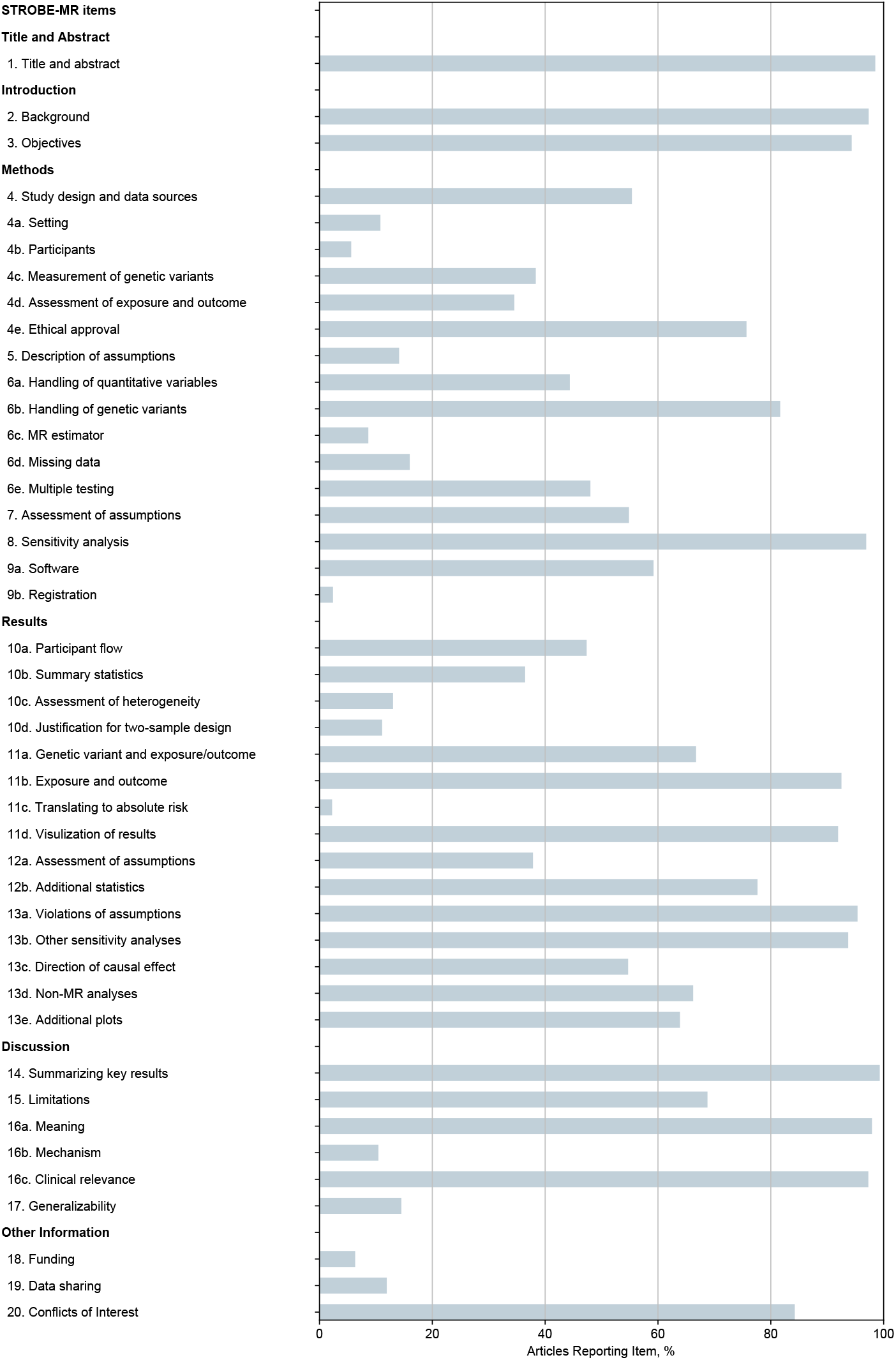
STROBE-MR reporting by item.

### 5.3 Reporting Trends in Different Sections

The reporting rate of STROBE-MR items varied significantly in different sections. As can be seen in Figure 2, items in Title/Abstract and Introduction sections were reported by most of the articles, with reporting rates of 98.5% and 95.8%, respectively. However, reporting rates decreased significantly in other sections: followed by Results (51.3%), Discussion (48.0%) and Methods (32.8%). The Other Information section had the lowest reporting rate of 21.2%. As can be seen in Table S1-6, only 24.5% articles reported the role of funders and only 12.1% shared statistical code. In the Methods section, item 4b: participants and item 6c: details of the MR estimator were rarely reported. In the Discussion section, item 17: generalizability and item 16b: Mechanism was most likely to be omitted by authors. Table S1-6 shows that the gene-environment equivalence component of item 16b only has a reporting rate of 10.0% and generalizability across other exposure periods and levels components of item 17 had reporting rates of 26.2% and 20.0%, respectively.

**Figure 2:**
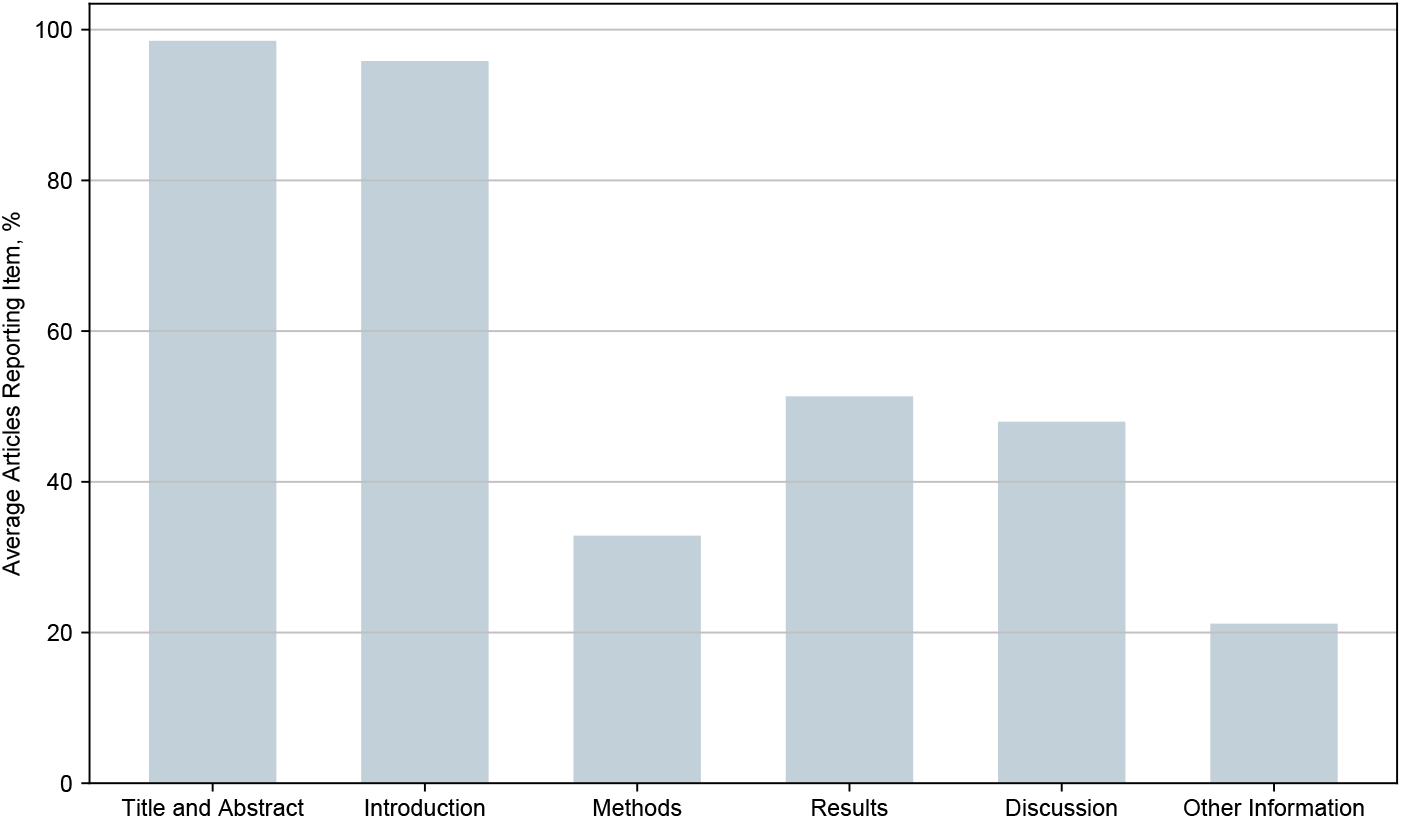
STROBE-MR reporting by section.

Different temporal trends in reporting can also be observed in these sections (Figure 3). The reporting rates remained around 90% in the Title/Abstract and Introduction sections. The Results section also showed a constant temporal trend but its reporting rate was only around 50%. The Methods, Discussion and Other Information sections all showed a decreasing trend, especially in recent years.

**Figure 3:**
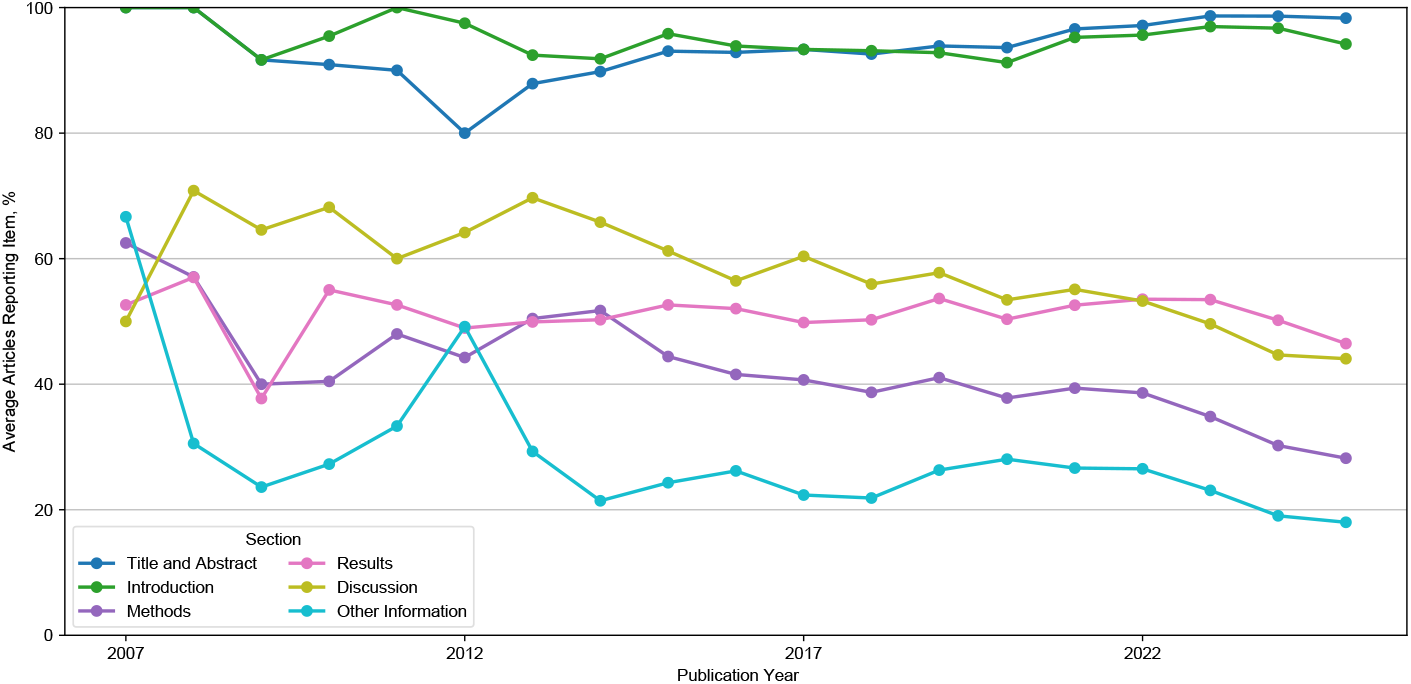
STROBE-MR reporting by year and section.

### 5.4 Sensitivity Analysis

Detailed results of the sensitivity analysis, which used a partial compliance criterion, can be found in the Appendix. Under the partial compliance criterion, the reporting rate of many items was higher and an average of 32 items met this criterion. Figure S2-1 shows there was a similar stable trend before 2021 and a decreasing trend after 2021. However, we observed significant changes in reporting rates of different items. The most frequently reported items were now item 4 (99.5%), 14 (99.3%), 1 (98.5%), 16b (98.5%) and 16a (98.0%). The least reported items were item 11c (2.2%), 9b (2.4%), 10c (13.0%), 6d (16.0%), 10b (36.5%). When comparing reporting rate in different sections, we observed a significantly higher reporting rate in the Methods (83.5%), Discussion (94.8%) and Other Information (86.7%) sections. These are the least reported sections in the main analysis.

Temporal trends were similar under the partial and complete compliance criteria in the Title/Abstract, Introduction and Results sections. Partial compliance remained stable in the Methods and Discussion sections, while complete compliance declined. Temporal patterns also diverged in the Other Information section: the reporting rate rose from around 60% to over 80% in recent years, contrasting with the declining trend in complete compliance.

## 6 Discussion

This study provides, to our knowledge, the first large-scale automated evaluation of STROBE-MR reporting quality in MR studies using an LLM. By analyzing more than 12,000 MR articles published between 2007 and 2025, several important patterns emerged regarding both the strengths and weaknesses of current reporting practices. Overall, MR studies demonstrate high compliance with some core STROBE-MR items, including research background, key results, and the main findings and interpretations. However, this contrasts with the poor reporting of several methodological and transparency-related items, particularly in the Other Information, Methods, and Discussion sections. Reporting quality has also not improved over time, even after the introduction of STROBE-MR.

Our results are consistent with previous studies that assessed MR reporting on a smaller scale. A recent systematic review [14] of MR studies on risk factors for vascular dementia found that only 10 of the 26 articles (38%) included a STROBE-MR checklist. Our findings also show that only 26.2% of articles published after 2021 cited STROBE-MR. This suggests that STROBE-MR is not widely accepted in the MR research community, and the interpretation of published MR research should therefore take into account this inconsistency in reporting. Across different STROBE-MR sections and items, a previous study [17] of 111 articles also identified low reporting rates for items in the Methods section, such as IV assumptions and details of MR estimators and covariates. One explanation for this poor reporting is the dominance of two-sample MR designs (88.6%), particularly in recent years. These articles rely on secondary data sources, which may lead authors to omit detailed descriptions of participants, data collection, and assumptions. This may be based on an assumption that these are already understood or previously reported, or may potentially be because the original data source did not provide sufficient detail. However, such omissions hinder transparency and reproducibility, especially for readers who are not domain experts. Similar low reporting rates can also be found in the Other Information section: 8% of MR articles provided statistical code in the study by Gibson et al. (12% in our study). Sharing statistical code is key to reproducing results, and a low sharing rate could affect the reproducibility of MR studies. The same study also found that reporting quality remained stable over time, which is consistent with our findings.

The sensitivity analysis using the partial compliance criterion showed substantially higher reporting rates in sections with low complete compliance rates in the main analysis. This indicates that many studies reported some, but not all, applicable components of these items. Interestingly, we found no difference in the average number of items reported between studies with and without self-claimed STROBE-MR compliance. These findings indicate that authors have different interpretations of STROBE-MR, and that the guideline was not applied in a consistent way.

Our findings also suggest potential refinements to the STROBE-MR guideline itself. The difference between complete and partial compliance rates highlights incomplete reporting of compound STROBE-MR items. Explicitly separating these components and clarifying which components are required or optional could help authors report more consistently. Given the predominance of two-sample designs, additional guidance tailored for two-sample studies could be helpful. For example, additional guidance could clarify which details of the data source (e.g., GWAS datasets) should be disclosed in the report and when references to the original data sources are sufficient. We also found that lower agreement with the human consensus in the Methods and Results sections coincided with greater disagreement between human reviewers in these sections, suggesting that disagreement with human consensus may partly reflect ambiguity in the assessment task itself, rather than limitations in LLM performance. Some technical items in these sections may benefit from clarification. For example, the LLM confused the measurement of genetic variants with the selection of genetic instruments. Further examples of cases in which the model differ from both reviewers or matched final human consensus despite disagreement between reviwers are presented in Supplementary Table **??**.

These findings suggest that the introduction of the STROBE-MR guideline has not yet translated into uniformly improved reporting practices across the MR literature. Although certain items are well reported, key methodological and transparency elements remain neglected. This may reflect limited awareness of the guideline, lack of enforcement by journals and reviewers, or misunderstanding of certain items. To address these issues, stronger endorsement of STROBE-MR by journals, peer reviewers, and funding bodies, along with more training on how to apply STROBE-MR, may be necessary.

### 6.1 Limitations

The strengths of this study include its scale and the comprehensiveness of the MR literature assessments. However, there are also several limitations. First, the reliance on available full text on PubMed Central may introduce time-varying selection bias as more publishers are joining the open-assess community. Earlier aricles on PubMed Central may reprensent authors that favored open science and thus are more inclined to transparent reporting. Second, the accuracy of LLM-based assessment depends on prompt design and model selection: we only used GPT-5-mini, and other models may achieve better or worse results. We also did not leverage the multimodal capabilities of the model because almost all information we needed for the assessment was available in text format. However, multimodality support is rapidly improving in frontier LLMs and could further enhance performance.

### 6.2 Implications

Our findings have implications for various parties. For the MR community, our results suggest that important methodological details are not consistently reported across different studies, and further efforts are needed to promote the use of STROBE-MR. For evidence synthesis researchers, our study provides a useful tool that can facilitate the rapid screening and identification of well-reported MR studies. Our tool can also be integrated into journal submission workflows to help editors identify reporting problems and provide authors with feedback. For reporting guideline researchers, our work demonstrates the potential of applying LLMs to large-scale reporting quality monitoring, which can then be used to improve the guideline itself. Finally, this study provides an automated reporting assessment workflow that can be adapted to other reporting guidelines.

## 7 Conclusion

In summary, MR studies generally report their background, aims, key results and findings clearly. Significant gaps remain in methodological transparency and reproducibility. Reporting quality has not improved over time and may even be declining in recent years, despite the availability of reporting guidelines. LLM-based approaches offer a promising tool for assessing and monitoring transparent reporting of medical literature at scale. Future efforts can focus on integrating LLM into literature publication workflows and educational intervention to increase engagement of MR researchers with STROBE-MR.

## Data and Code availability

The evaluation dataset and code for analysis are available at the following GitHub repository: https://github.com/ActionSafe/strobe-mr-code. The annotation tool is available at: https://github.com/ActionSafe/AnnotationToolMR.

## Footnotes

1 https://developers.openai.com/api/reference/resources/responses/methods/create

## References

[1] Smith GD, Ebrahim S. ‘Mendelian randomization’: can genetic epidemiology contribute to understanding environmental determinants of disease? International Journal of Epidemiology. 2003;32(1):1–22.

[2] Nguyen K, Mitchell BD. A Guide to Understanding Mendelian Randomization Studies. Arthritis Care Research. 2024 Aug;76(11):1451–1460. Available from: 10.1002/acr.25400.

[3] Davies NM, Holmes MV, Davey Smith G. Reading Mendelian randomisation studies: a guide, glossary, and checklist for clinicians. BMJ. 2018 Jul:k601. Available from: 10.1136/bmj.k601.

[4] Burgess S, Thompson SG. Multivariable Mendelian Randomization: The Use of Pleiotropic Genetic Variants to Estimate Causal Effects. American Journal of Epidemiology. 2015 Jan;181(4):251–260. Available from: 10.1093/aje/kwu283.

[5] Burgess S, Davey Smith G, Davies NM, Dudbridge F, Gill D, Glymour MM, et al. Guidelines for performing Mendelian randomization investigations: update for summer 2023. Wellcome Open Research. 2023 Aug;4:186. Available from: https://pmc.ncbi.nlm.nih.gov/articles/PMC7384151/.

[6] Burgess S, Daniel RM, Butterworth AS, Thompson SG. Network Mendelian randomization: using genetic variants as instrumental variables to investigate mediation in causal pathways. International Journal of Epidemiology. 2015 Apr;44(2):484–95. Available from: https://pmc.ncbi.nlm.nih.gov/articles/PMC4469795/.

[7] Hemani G, Zheng J, Elsworth B, Wade KH, Haberland V, Baird D, et al. The MR-Base platform supports systematic causal inference across the human phenome. eLife. 2018 May;7:e34408. Available from: 10.7554/eLife.34408.

[8] Sudlow C, Gallacher J, Allen N, Beral V, Burton P, Danesh J, et al. UK biobank: an open access resource for identifying the causes of a wide range of complex diseases of middle and old age. PLoS medicine. 2015 Mar;12(3):e1001779.

[9] He K, Wu X, Qian Z, Lin K, Wang Y, Su Z, et al. The rise of Mendelian randomization for exploring disease causality: A bibliometric analysis based on CiteSpace. Medicine. 2025 Aug;104(32):e43764. Available from: 10.1097/MD.0000000000043764.

[10] Skrivankova VW, Richmond RC, Woolf BAR, Yarmolinsky J, Davies NM, Swanson SA, et al. Strengthening the Reporting of Observational Studies in Epidemiology Using Mendelian Randomization: The STROBE-MR Statement. JAMA. 2021;326(16):1614–21.

[11] Davies NM, Smith GD, Windmeijer F, Martin RM. Issues in the reporting and conduct of instrumental variable studies: a systematic review. Epidemiology. 2013;24(3):363–9.

[12] Islam SN, Ahammed T, Anjum A, Albalawi O, Uddin MJ. Reporting methodological issues of the mendelian randomization studies in health and medical research: a systematic review. BMC medical research methodology. 2022;22(1):21.

[13] VanderWeele TJ, Tchetgen Tchetgen EJ, Cornelis M, Kraft P. Methodological Challenges in Mendelian Randomization. Epidemiology. 2014 May;25(3):427–435. Available from: 10.1097/EDE.0000000000000081.

[14] Zuo M, Gagliano Taliun SA. A systematic review of risk factors for vascular dementia using Mendelian randomization: a call for methodologically robust studies to advance knowledge. BMC neurology. 2026 Apr;26(1):359.

[15] Cui Y, Xu J, Zheng Q, Wang J, Xu C, Zhang M, et al. Genetically proxied gut microbiota and cancer risk: a scoping review of Mendelian randomization studies. Archives of Public Health. 2025 Aug;83(1):209. Available from: 10.1186/s13690-025-01685-2.

[16] Li X, Liu Q, Ni H, Ni J, Yang S, Ji J. Causal factors for migraine in Mendelian randomization studies: a systematic review and meta-analysis. Frontiers in Neurology. 2025;16:1660995. Available from: https://pmc.ncbi.nlm.nih.gov/articles/PMC12454037/.

[17] Gibson MJ, Spiga F, Campbell A, Khouja JN, Richmond RC, Munafò MR. Reporting and methodological quality of studies that use Mendelian randomisation in UK Biobank: a meta-epidemiological study. BMJ Evidence-Based Medicine. 2023 Apr;28(2):103–10. Available from: https://pmc.ncbi.nlm.nih.gov/articles/PMC10086297/.

[18] Legate A, Nimon K, Noblin A. (Semi)automated approaches to data extraction for systematic reviews and meta-analyses in social sciences: A living review. F1000Research. 2024 Sep;13:664. Available from: https://pmc.ncbi.nlm.nih.gov/articles/PMC11364972/.

[19] Laignelot F, Martin GL, Ossman M, Pingeon O, Boubaker A, Picovschi E, et al. Large language models show promising performance for some systematic review tasks but call for cautious implementation: a systematic review. Journal of Clinical Epidemiology. 2026 Mar;194:112221.

[20] Nyrhi L, Ponkilainen V, Laaksonen J, Kuikka L, Paljakka L, Karjalainen T, et al. Large language models for risk-of-bias assessment in randomised clinical trials—a comparative validation study. eBioMedicine. 2026 Mar;126:106238. Available from: https://pmc.ncbi.nlm.nih.gov/articles/PMC13054287/.

[21] Lai H, Ge L, Sun M, Pan B, Huang J, Hou L, et al. Assessing the Risk of Bias in Randomized Clinical Trials With Large Language Models. JAMA Network Open. 2024 May;7(5):e2412687. Available from: 10.1001/jamanetworkopen.2024.12687.

[22] Srinivasan A, Berkowitz J, Friedrich NA, Kivelson S, Tatonetti NP. Large Language Model Analysis of Reporting Quality of Randomized Clinical Trial Articles: A Systematic Review. JAMA Network Open. 2025 Aug;8(8):e2529418. Available from: 10.1001/jamanetworkopen.2025.29418.

[23] Luo X, Li Z, Yang Z, Wang B, Ma Y, Chen F, et al. Using Large Language Models to Assess the Consistency of Randomized Controlled Trials on AI Interventions With CONSORT-AI: Cross-Sectional Survey. Journal of Medical Internet Research. 2025 Sep;27(1):e72412. Available from: https://www.jmir.org/2025/1/e72412.

