## Supplementary materials for "Reporting Quality of Mendelian Randomization Studies Based on STROBE-MR: An Investigation Powered by Large Language Models"

2026-10-21

### S1 Supplementary tables

Table S1-1: Definitions of STROBE-MR components

| Index | Section | Item | Component |
|---|---|---|---|
| 1 | Title and Abstract | 1 | Indicate Mendelian randomization (MR) as the study's design in the title and/or the abstract if that is a main purpose of the study |
| 2 | Introduction | 2 | Explain the scientific background and rationale for the reported study. What is the exposure? Is a potential causal relationship between exposure and outcome plausible? Justify why MR is a helpful method to address the study question |
| 3 | Introduction | 3 | State specific objectives clearly, including pre-specified causal hypotheses (if any). State that MR is a method that, under specific assumptions, intends to estimate causal effects |
| 4 | Methods | 4 | Present data sources of the study early in the article |
| 5 | Methods | 4 | Present 2. Methods of the study in the article |
| 6 | Methods | 4 | Present 3. Exposures of the study in the article |
| 7 | Methods | 4 | Present 4. Outcomes of the study in the article |
| 8 | Methods | 4 | including a table listing sources of data for all phases of the study |
| 9 | Methods | 4a | [Setting] For each data source, describe its study design |
| 10 | Methods | 4a | [Setting] For each data source, describe its underlying population |
| 11 | Methods | 4a | [Setting] For each data source, describe its locations |
| 12 | Methods | 4a | [Setting] For each data source, describe relevant dates, including periods of recruitment |
| 13 | Methods | 4a | [Setting] For each data source, describe its exposure |
| 14 | Methods | 4a | [Setting] For each data source, describe how the follow-up process was conducted |
| 15 | Methods | 4a | [Setting] For each data source, describe the data collection process |
| 16 | Methods | 4b | [Participants] For each data source, give the eligibility criteria, and the sources and methods of selection of participants |
| 17 | Methods | 4b | [Participants] For each data source, report the sample size |
| 18 | Methods | 4b | [Participants] For each data source, report whether any power or sample size calculations were carried out prior to the main analysis |
| 19 | Methods | 4c | [Participants] For each data source, describe measurement of genetic variants |
| 20 | Methods | 4c | [Participants] For each data source, describe quality control of genetic variants |
| 21 | Methods | 4c | [Participants] For each data source, describe selection of genetic variants |
| 22 | Methods | 4d | Describe methods of assessment and diagnostic criteria for each exposure |
| 23 | Methods | 4d | Describe methods of assessment and diagnostic criteria for each outcome |
| 24 | Methods | 4d | Describe methods of assessment and diagnostic criteria for other relevant variables |
| 25 | Methods | 4e | Provide details of ethics committee approval and participant informed consent, if relevant |
| 26 | Methods | 5 | Explicitly state the relevance assumption of IV |
| 27 | Methods | 5 | Explicitly state the independence assumption of IV |
| 28 | Methods | 5 | Explicitly state the exclusion assumption of IV |
| 29 | Methods | 5 | State assumptions for any additional or sensitivity analysis |
| 30 | Methods | 6a | Describe how quantitative variables were handled in the analyses (i.e., scale, units, model) |
| 31 | Methods | 6b | Describe how genetic variants were handled in the analyses and, if applicable, how their weights were selected |
| 32 | Methods | 6c | Describe the MR estimator (e.g. two-stage least squares, Wald ratio) and related statistics. |
| 33 | Methods | 6c | Detail the included covariates |
| 34 | Methods | 6c | in case of two-sample MR, whether the same covariate set was used for adjustment in the two samples |
| 35 | Methods | 6d | Explain how missing data were addressed |

| Index | Section | Item | Component |
|---|---|---|---|
| 36 | Methods | 6e | If applicable, indicate how multiple testing was addressed |
| 37 | Methods | 7 | Describe any methods or prior knowledge used to assess the relevance assumption of IV |
| 38 | Methods | 7 | Describe any methods or prior knowledge used to assess the independence assumption of IV |
| 39 | Methods | 7 | Describe any methods or prior knowledge used to assess the exclusion assumption of IV |
| 40 | Methods | 7 | Describe any methods or prior knowledge used to assess the assumptions for any additional analysis or sensitivity analysis |
| 41 | Methods | 8 | Describe any sensitivity analyses or additional analyses performed |
| 42 | Methods | 9a | Name statistical software and package(s), including version and settings used |
| 43 | Methods | 9b | State whether the study protocol and details were pre-registered (as well as when and where) |
| 44 | Results | 10a | Report the numbers of individuals at each stage of included studies and reasons for exclusion. Consider use of a flow diagram |
| 45 | Results | 10b | Report summary statistics for phenotypic exposure(s), outcome(s), and other relevant variables (e.g. means, SDs, proportions) |
| 46 | Results | 10c | If the data sources include meta-analyses of previous studies, provide the assessments of heterogeneity across these studies |
| 47 | Results | 10d | For two-sample MR: Provide justification of the similarity of the genetic variant-exposure associations between the exposure and outcome samples |
| 48 | Results | 10d | For two-sample MR: Provide information on the number of individuals who overlap between the exposure and outcome studies |
| 49 | Results | 11a | Report the associations between genetic variant and exposure, and between genetic variant and outcome, preferably on an interpretable scale |
| 50 | Results | 11b | Report MR estimates of the relationship between exposure and outcome, and the measures of uncertainty from the MR analysis, on an interpretable scale, such as odds ratio or relative risk per SD difference |
| 51 | Results | 11c | Describe whether translating estimates of relative risk into absolute risk is conducted |
| 52 | Results | 11d | Consider plots to visualize results (e.g. forest plot, scatterplot of associations between genetic variants and outcome versus between genetic variants and exposure) |
| 53 | Results | 12a | Report results of the assessment of the relevance assumption |
| 54 | Results | 12a | Report results of the assessment of the independence assumption |
| 55 | Results | 12a | Report results of the assessment of the exclusion assumption |
| 56 | Results | 12a | Report results of the assessment of the any additional assumption, if relevant |
| 57 | Results | 12b | Report any additional statistics |
| 58 | Results | 13a | Report results from any sensitivity analyses to assess the robustness of the main results to violations of the assumptions |
| 59 | Results | 13b | Report results from other sensitivity analyses or additional analyses |
| 60 | Results | 13c | Report any assessment of direction of causal relationship (e.g., bidirectional MR) |
| 61 | Results | 13d | Report and compare with estimates from non-MR analyses |
| 62 | Results | 13e | Consider additional plots to visualize results (e.g., leave-one-out analyses) |
| 63 | Discussion | 14 | Summarize key results with reference to study objectives |
| 64 | Discussion | 15 | Limitation: discuss the validity of the IV assumptions |
| 65 | Discussion | 15 | Limitation: discuss other sources of potential bias and imprecision |
| 66 | Discussion | 15 | Limitation: discuss both direction and magnitude of any potential bias and any efforts to address them |
| 67 | Discussion | 16a | Meaning: Give a cautious overall interpretation of results in the context of their limitations and in comparison with other studies |
| 68 | Discussion | 16b | Mechanism: Discuss underlying biological mechanisms that could drive a potential causal relationship between the investigated exposure and the outcome |

| Index | Section | Item | Component |
|---|---|---|---|
| 69 | Discussion | 16b | Mechanism: Discuss whether the gene-environment equivalence assumption is reasonable |
| 70 | Discussion | 16b | Mechanism: Clarifying that IV estimates may provide causal effects only under certain assumptions |
| 71 | Discussion | 16c | Clinical relevance: Discuss whether the results have clinical or public policy relevance, and to what extent they inform effect sizes of possible interventions |
| 72 | Discussion | 17 | Discuss the generalizability of the study results to other populations |
| 73 | Discussion | 17 | Discuss the generalizability of the study results across other exposure periods/timings |
| 74 | Discussion | 17 | Discuss the generalizability of the study results across other levels of exposure |
| 75 | Other Information | 18 | Describe sources of funding in the present study |
| 76 | Other Information | 18 | Describe the role of funders in the present study |
| 77 | Other Information | 18 | If applicable, describe sources of funding for the databases or studies on which the present study is based |
| 78 | Other Information | 19 | Provide the data used to perform all analyses or report where and how the data can be accessed, and reference these sources in the article |
| 79 | Other Information | 19 | Provide the statistical code needed to reproduce the results in the article, or report whether the code is publicly accessible and if so, where |
| 80 | Other Information | 20 | All authors should declare all potential conflicts of interest |

Table S1-2: API parameters for OpenAI platform

| Parameter | Value |
|---|---|
| text_format | `class Assessment(BaseModel):`<br>` index: int`<br>` rationale: str`<br>` relevant_text: str = ""`<br>` status: int` |
| reasoning_effort | ”high” |

Table S1-3: Definition of JSON object

| Field | Explanation |
|---|---|
| pmcid | PMCID |
| title | title of the article |
| year | publication year |
| abstract | abstract of the article |
| keyword | keywords of the article |
| sections | sections in the main text |
| figures | links of figures included in the main text |
| attachments | links of attachment files |

Table S1-4: Annotation summary by component

| Index | Section | Item | JZ | | | XW | | | Consensus | | | Disagreement | Total |
|---|---|---|---|---|---|---|---|---|---|---|---|---|---|
| | | | Reported | NR | NA | Reported | NR | NA | Reported | NR | NA | | |
| 1 | Title and Abstract | 1 | 46 (100.0%) | 0 (0.0%) | 0 (0.0%) | 46 (100.0%) | 0 (0.0%) | 0 (0.0%) | 46 (100.0%) | 0 (0.0%) | 0 (0.0%) | 0 (0.0%) | 46 |
| 2 | Introduction | 2 | 46 (100.0%) | 0 (0.0%) | 0 (0.0%) | 46 (100.0%) | 0 (0.0%) | 0 (0.0%) | 46 (100.0%) | 0 (0.0%) | 0 (0.0%) | 0 (0.0%) | 46 |
| 3 | Introduction | 3 | 42 (91.3%) | 4 (8.7%) | 0 (0.0%) | 43 (93.5%) | 3 (6.5%) | 0 (0.0%) | 42 (91.3%) | 4 (8.7%) | 0 (0.0%) | 1 (2.2%) | 46 |
| 4 | Methods | 4 | 46 (100.0%) | 0 (0.0%) | 0 (0.0%) | 46 (100.0%) | 0 (0.0%) | 0 (0.0%) | 46 (100.0%) | 0 (0.0%) | 0 (0.0%) | 0 (0.0%) | 46 |
| 5 | Methods | 4 | 46 (100.0%) | 0 (0.0%) | 0 (0.0%) | 46 (100.0%) | 0 (0.0%) | 0 (0.0%) | 46 (100.0%) | 0 (0.0%) | 0 (0.0%) | 0 (0.0%) | 46 |
| 6 | Methods | 4 | 46 (100.0%) | 0 (0.0%) | 0 (0.0%) | 46 (100.0%) | 0 (0.0%) | 0 (0.0%) | 46 (100.0%) | 0 (0.0%) | 0 (0.0%) | 0 (0.0%) | 46 |
| 7 | Methods | 4 | 46 (100.0%) | 0 (0.0%) | 0 (0.0%) | 46 (100.0%) | 0 (0.0%) | 0 (0.0%) | 45 (97.8%) | 1 (2.2%) | 0 (0.0%) | 0 (0.0%) | 46 |

| Index | Section | Item | JZ | | | XW | | | Consensus | | | Disagreement | Total |
|---|---|---|---|---|---|---|---|---|---|---|---|---|---|
| | | | Reported | NR | NA | Reported | NR | NA | Reported | NR | NA | | |
| 8 | Methods | 4 | 27 (58.7%) | 19 (41.3%) | 0 (0.0%) | 27 (58.7%) | 19 (41.3%) | 0 (0.0%) | 28 (60.9%) | 18 (39.1%) | 0 (0.0%) | 2 (4.3%) | 46 |
| 9 | Methods | 4a | 24 (52.2%) | 22 (47.8%) | 0 (0.0%) | 28 (60.9%) | 18 (39.1%) | 0 (0.0%) | 26 (56.5%) | 20 (43.5%) | 0 (0.0%) | 6 (13.0%) | 46 |
| 10 | Methods | 4a | 36 (78.3%) | 10 (21.7%) | 0 (0.0%) | 39 (84.8%) | 7 (15.2%) | 0 (0.0%) | 40 (87.0%) | 6 (13.0%) | 0 (0.0%) | 7 (15.2%) | 46 |
| 11 | Methods | 4a | 26 (56.5%) | 20 (43.5%) | 0 (0.0%) | 23 (50.0%) | 23 (50.0%) | 0 (0.0%) | 33 (71.7%) | 13 (28.3%) | 0 (0.0%) | 9 (19.6%) | 46 |
| 12 | Methods | 4a | 11 (23.9%) | 35 (76.1%) | 0 (0.0%) | 7 (15.2%) | 39 (84.8%) | 0 (0.0%) | 9 (19.6%) | 37 (80.4%) | 0 (0.0%) | 4 (8.7%) | 46 |
| 13 | Methods | 4a | 38 (82.6%) | 8 (17.4%) | 0 (0.0%) | 36 (78.3%) | 10 (21.7%) | 0 (0.0%) | 35 (76.1%) | 11 (23.9%) | 0 (0.0%) | 4 (8.7%) | 46 |
| 14 | Methods | 4a | 5 (10.9%) | 32 (69.6%) | 9 (19.6%) | 6 (13.0%) | 21 (45.7%) | 19 (41.3%) | 6 (13.0%) | 26 (56.5%) | 14 (30.4%) | 18 (39.1%) | 46 |
| 15 | Methods | 4a | 16 (34.8%) | 30 (65.2%) | 0 (0.0%) | 19 (41.3%) | 27 (58.7%) | 0 (0.0%) | 24 (52.2%) | 22 (47.8%) | 0 (0.0%) | 9 (19.6%) | 46 |
| 16 | Methods | 4b | 10 (21.7%) | 36 (78.3%) | 0 (0.0%) | 10 (21.7%) | 36 (78.3%) | 0 (0.0%) | 11 (23.9%) | 35 (76.1%) | 0 (0.0%) | 2 (4.3%) | 46 |
| 17 | Methods | 4b | 43 (93.5%) | 3 (6.5%) | 0 (0.0%) | 44 (95.7%) | 2 (4.3%) | 0 (0.0%) | 43 (93.5%) | 3 (6.5%) | 0 (0.0%) | 1 (2.2%) | 46 |
| 18 | Methods | 4b | 4 (8.7%) | 42 (91.3%) | 0 (0.0%) | 7 (15.2%) | 39 (84.8%) | 0 (0.0%) | 4 (8.7%) | 42 (91.3%) | 0 (0.0%) | 3 (6.5%) | 46 |
| 19 | Methods | 4c | 16 (34.8%) | 30 (65.2%) | 0 (0.0%) | 13 (28.3%) | 32 (69.6%) | 1 (2.2%) | 27 (58.7%) | 19 (41.3%) | 0 (0.0%) | 12 (26.1%) | 46 |
| 20 | Methods | 4c | 16 (34.8%) | 30 (65.2%) | 0 (0.0%) | 11 (23.9%) | 35 (76.1%) | 0 (0.0%) | 32 (69.6%) | 14 (30.4%) | 0 (0.0%) | 15 (32.6%) | 46 |
| 21 | Methods | 4c | 46 (100.0%) | 0 (0.0%) | 0 (0.0%) | 46 (100.0%) | 0 (0.0%) | 0 (0.0%) | 46 (100.0%) | 0 (0.0%) | 0 (0.0%) | 0 (0.0%) | 46 |
| 22 | Methods | 4d | 29 (63.0%) | 17 (37.0%) | 0 (0.0%) | 28 (60.9%) | 18 (39.1%) | 0 (0.0%) | 29 (63.0%) | 17 (37.0%) | 0 (0.0%) | 1 (2.2%) | 46 |
| 23 | Methods | 4d | 24 (52.2%) | 22 (47.8%) | 0 (0.0%) | 26 (56.5%) | 20 (43.5%) | 0 (0.0%) | 27 (58.7%) | 19 (41.3%) | 0 (0.0%) | 4 (8.7%) | 46 |
| 24 | Methods | 4d | 13 (28.3%) | 33 (71.7%) | 0 (0.0%) | 16 (34.8%) | 30 (65.2%) | 0 (0.0%) | 16 (34.8%) | 30 (65.2%) | 0 (0.0%) | 5 (10.9%) | 46 |
| 25 | Methods | 4e | 40 (87.0%) | 5 (10.9%) | 1 (2.2%) | 41 (89.1%) | 4 (8.7%) | 1 (2.2%) | 42 (91.3%) | 3 (6.5%) | 1 (2.2%) | 6 (13.0%) | 46 |
| 26 | Methods | 5 | 32 (69.6%) | 14 (30.4%) | 0 (0.0%) | 37 (80.4%) | 9 (19.6%) | 0 (0.0%) | 39 (84.8%) | 7 (15.2%) | 0 (0.0%) | 5 (10.9%) | 46 |
| 27 | Methods | 5 | 32 (69.6%) | 14 (30.4%) | 0 (0.0%) | 35 (76.1%) | 11 (23.9%) | 0 (0.0%) | 37 (80.4%) | 9 (19.6%) | 0 (0.0%) | 3 (6.5%) | 46 |
| 28 | Methods | 5 | 32 (69.6%) | 14 (30.4%) | 0 (0.0%) | 35 (76.1%) | 11 (23.9%) | 0 (0.0%) | 37 (80.4%) | 9 (19.6%) | 0 (0.0%) | 3 (6.5%) | 46 |
| 29 | Methods | 5 | 8 (17.4%) | 38 (82.6%) | 0 (0.0%) | 8 (17.4%) | 38 (82.6%) | 0 (0.0%) | 7 (15.2%) | 39 (84.8%) | 0 (0.0%) | 2 (4.3%) | 46 |
| 30 | Methods | 6a | 19 (41.3%) | 23 (50.0%) | 4 (8.7%) | 16 (34.8%) | 30 (65.2%) | 0 (0.0%) | 18 (39.1%) | 28 (60.9%) | 0 (0.0%) | 7 (15.2%) | 46 |
| 31 | Methods | 6b | 44 (95.7%) | 2 (4.3%) | 0 (0.0%) | 42 (91.3%) | 4 (8.7%) | 0 (0.0%) | 44 (95.7%) | 2 (4.3%) | 0 (0.0%) | 2 (4.3%) | 46 |
| 32 | Methods | 6c | 46 (100.0%) | 0 (0.0%) | 0 (0.0%) | 46 (100.0%) | 0 (0.0%) | 0 (0.0%) | 46 (100.0%) | 0 (0.0%) | 0 (0.0%) | 0 (0.0%) | 46 |
| 33 | Methods | 6c | 15 (32.6%) | 31 (67.4%) | 0 (0.0%) | 15 (32.6%) | 31 (67.4%) | 0 (0.0%) | 14 (30.4%) | 32 (69.6%) | 0 (0.0%) | 2 (4.3%) | 46 |
| 34 | Methods | 6c | 3 (6.5%) | 40 (87.0%) | 3 (6.5%) | 2 (4.3%) | 41 (89.1%) | 3 (6.5%) | 3 (6.5%) | 40 (87.0%) | 3 (6.5%) | 1 (2.2%) | 46 |
| 35 | Methods | 6d | 5 (10.9%) | 41 (89.1%) | 0 (0.0%) | 6 (13.0%) | 40 (87.0%) | 0 (0.0%) | 8 (17.4%) | 38 (82.6%) | 0 (0.0%) | 5 (10.9%) | 46 |
| 36 | Methods | 6e | 24 (52.2%) | 22 (47.8%) | 0 (0.0%) | 23 (50.0%) | 23 (50.0%) | 0 (0.0%) | 23 (50.0%) | 23 (50.0%) | 0 (0.0%) | 1 (2.2%) | 46 |
| 37 | Methods | 7 | 42 (91.3%) | 4 (8.7%) | 0 (0.0%) | 41 (89.1%) | 5 (10.9%) | 0 (0.0%) | 42 (91.3%) | 4 (8.7%) | 0 (0.0%) | 1 (2.2%) | 46 |
| 38 | Methods | 7 | 24 (52.2%) | 22 (47.8%) | 0 (0.0%) | 30 (65.2%) | 16 (34.8%) | 0 (0.0%) | 28 (60.9%) | 18 (39.1%) | 0 (0.0%) | 6 (13.0%) | 46 |
| 39 | Methods | 7 | 43 (93.5%) | 3 (6.5%) | 0 (0.0%) | 44 (95.7%) | 2 (4.3%) | 0 (0.0%) | 44 (95.7%) | 2 (4.3%) | 0 (0.0%) | 3 (6.5%) | 46 |
| 40 | Methods | 7 | 18 (39.1%) | 28 (60.9%) | 0 (0.0%) | 38 (82.6%) | 8 (17.4%) | 0 (0.0%) | 28 (60.9%) | 18 (39.1%) | 0 (0.0%) | 20 (43.5%) | 46 |
| 41 | Methods | 8 | 46 (100.0%) | 0 (0.0%) | 0 (0.0%) | 46 (100.0%) | 0 (0.0%) | 0 (0.0%) | 46 (100.0%) | 0 (0.0%) | 0 (0.0%) | 0 (0.0%) | 46 |
| 42 | Methods | 9a | 23 (50.0%) | 23 (50.0%) | 0 (0.0%) | 19 (41.3%) | 27 (58.7%) | 0 (0.0%) | 24 (52.2%) | 22 (47.8%) | 0 (0.0%) | 14 (30.4%) | 46 |
| 43 | Methods | 9b | 3 (6.5%) | 43 (93.5%) | 0 (0.0%) | 3 (6.5%) | 43 (93.5%) | 0 (0.0%) | 3 (6.5%) | 43 (93.5%) | 0 (0.0%) | 0 (0.0%) | 46 |
| 44 | Results | 10a | 14 (30.4%) | 31 (67.4%) | 1 (2.2%) | 16 (34.8%) | 29 (63.0%) | 1 (2.2%) | 16 (34.8%) | 29 (63.0%) | 1 (2.2%) | 2 (4.3%) | 46 |
| 45 | Results | 10b | 16 (34.8%) | 30 (65.2%) | 0 (0.0%) | 18 (39.1%) | 28 (60.9%) | 0 (0.0%) | 15 (32.6%) | 31 (67.4%) | 0 (0.0%) | 6 (13.0%) | 46 |
| 46 | Results | 10c | 4 (8.7%) | 17 (37.0%) | 25 (54.3%) | 5 (10.9%) | 31 (67.4%) | 10 (21.7%) | 5 (10.9%) | 26 (56.5%) | 15 (32.6%) | 16 (34.8%) | 46 |
| 47 | Results | 10d | 19 (41.3%) | 24 (52.2%) | 3 (6.5%) | 12 (26.1%) | 31 (67.4%) | 3 (6.5%) | 14 (30.4%) | 29 (63.0%) | 3 (6.5%) | 7 (15.2%) | 46 |
| 48 | Results | 10d | 6 (13.0%) | 37 (80.4%) | 3 (6.5%) | 6 (13.0%) | 37 (80.4%) | 3 (6.5%) | 6 (13.0%) | 37 (80.4%) | 3 (6.5%) | 0 (0.0%) | 46 |
| 49 | Results | 11a | 33 (71.7%) | 13 (28.3%) | 0 (0.0%) | 33 (71.7%) | 13 (28.3%) | 0 (0.0%) | 34 (73.9%) | 12 (26.1%) | 0 (0.0%) | 2 (4.3%) | 46 |
| 50 | Results | 11b | 45 (97.8%) | 1 (2.2%) | 0 (0.0%) | 46 (100.0%) | 0 (0.0%) | 0 (0.0%) | 46 (100.0%) | 0 (0.0%) | 0 (0.0%) | 1 (2.2%) | 46 |
| 51 | Results | 11c | 0 (0.0%) | 46 (100.0%) | 0 (0.0%) | 0 (0.0%) | 45 (97.8%) | 1 (2.2%) | 0 (0.0%) | 45 (97.8%) | 1 (2.2%) | 1 (2.2%) | 46 |
| 52 | Results | 11d | 45 (97.8%) | 1 (2.2%) | 0 (0.0%) | 46 (100.0%) | 0 (0.0%) | 0 (0.0%) | 46 (100.0%) | 0 (0.0%) | 0 (0.0%) | 1 (2.2%) | 46 |
| 53 | Results | 12a | 37 (80.4%) | 9 (19.6%) | 0 (0.0%) | 36 (78.3%) | 10 (21.7%) | 0 (0.0%) | 36 (78.3%) | 10 (21.7%) | 0 (0.0%) | 3 (6.5%) | 46 |
| 54 | Results | 12a | 24 (52.2%) | 22 (47.8%) | 0 (0.0%) | 32 (69.6%) | 14 (30.4%) | 0 (0.0%) | 27 (58.7%) | 19 (41.3%) | 0 (0.0%) | 8 (17.4%) | 46 |
| 55 | Results | 12a | 43 (93.5%) | 3 (6.5%) | 0 (0.0%) | 46 (100.0%) | 0 (0.0%) | 0 (0.0%) | 45 (97.8%) | 1 (2.2%) | 0 (0.0%) | 3 (6.5%) | 46 |
| 56 | Results | 12a | 16 (34.8%) | 30 (65.2%) | 0 (0.0%) | 26 (56.5%) | 20 (43.5%) | 0 (0.0%) | 20 (43.5%) | 26 (56.5%) | 0 (0.0%) | 10 (21.7%) | 46 |
| 57 | Results | 12b | 42 (91.3%) | 4 (8.7%) | 0 (0.0%) | 42 (91.3%) | 4 (8.7%) | 0 (0.0%) | 42 (91.3%) | 4 (8.7%) | 0 (0.0%) | 0 (0.0%) | 46 |
| 58 | Results | 13a | 45 (97.8%) | 1 (2.2%) | 0 (0.0%) | 46 (100.0%) | 0 (0.0%) | 0 (0.0%) | 46 (100.0%) | 0 (0.0%) | 0 (0.0%) | 1 (2.2%) | 46 |
| 59 | Results | 13b | 44 (95.7%) | 2 (4.3%) | 0 (0.0%) | 43 (93.5%) | 3 (6.5%) | 0 (0.0%) | 45 (97.8%) | 1 (2.2%) | 0 (0.0%) | 3 (6.5%) | 46 |
| 60 | Results | 13c | 17 (37.0%) | 28 (60.9%) | 1 (2.2%) | 17 (37.0%) | 27 (58.7%) | 2 (4.3%) | 17 (37.0%) | 26 (56.5%) | 3 (6.5%) | 1 (2.2%) | 46 |
| 61 | Results | 13d | 19 (41.3%) | 27 (58.7%) | 0 (0.0%) | 15 (32.6%) | 30 (65.2%) | 1 (2.2%) | 29 (63.0%) | 17 (37.0%) | 0 (0.0%) | 17 (37.0%) | 46 |
| 62 | Results | 13e | 35 (76.1%) | 11 (23.9%) | 0 (0.0%) | 30 (65.2%) | 16 (34.8%) | 0 (0.0%) | 35 (76.1%) | 11 (23.9%) | 0 (0.0%) | 7 (15.2%) | 46 |
| 63 | Discussion | 14 | 46 (100.0%) | 0 (0.0%) | 0 (0.0%) | 46 (100.0%) | 0 (0.0%) | 0 (0.0%) | 46 (100.0%) | 0 (0.0%) | 0 (0.0%) | 0 (0.0%) | 46 |
| 64 | Discussion | 15 | 37 (80.4%) | 9 (19.6%) | 0 (0.0%) | 40 (87.0%) | 6 (13.0%) | 0 (0.0%) | 41 (89.1%) | 5 (10.9%) | 0 (0.0%) | 3 (6.5%) | 46 |
| 65 | Discussion | 15 | 45 (97.8%) | 1 (2.2%) | 0 (0.0%) | 44 (95.7%) | 2 (4.3%) | 0 (0.0%) | 45 (97.8%) | 1 (2.2%) | 0 (0.0%) | 1 (2.2%) | 46 |
| 66 | Discussion | 15 | 30 (65.2%) | 16 (34.8%) | 0 (0.0%) | 36 (78.3%) | 10 (21.7%) | 0 (0.0%) | 35 (76.1%) | 11 (23.9%) | 0 (0.0%) | 6 (13.0%) | 46 |
| 67 | Discussion | 16a | 46 (100.0%) | 0 (0.0%) | 0 (0.0%) | 46 (100.0%) | 0 (0.0%) | 0 (0.0%) | 46 (100.0%) | 0 (0.0%) | 0 (0.0%) | 0 (0.0%) | 46 |
| 68 | Discussion | 16b | 46 (100.0%) | 0 (0.0%) | 0 (0.0%) | 46 (100.0%) | 0 (0.0%) | 0 (0.0%) | 46 (100.0%) | 0 (0.0%) | 0 (0.0%) | 0 (0.0%) | 46 |
| 69 | Discussion | 16b | 9 (19.6%) | 37 (80.4%) | 0 (0.0%) | 10 (21.7%) | 36 (78.3%) | 0 (0.0%) | 11 (23.9%) | 35 (76.1%) | 0 (0.0%) | 1 (2.2%) | 46 |
| 70 | Discussion | 16b | 40 (87.0%) | 6 (13.0%) | 0 (0.0%) | 42 (91.3%) | 4 (8.7%) | 0 (0.0%) | 41 (89.1%) | 5 (10.9%) | 0 (0.0%) | 2 (4.3%) | 46 |
| 71 | Discussion | 16c | 46 (100.0%) | 0 (0.0%) | 0 (0.0%) | 46 (100.0%) | 0 (0.0%) | 0 (0.0%) | 46 (100.0%) | 0 (0.0%) | 0 (0.0%) | 0 (0.0%) | 46 |
| 72 | Discussion | 17 | 41 (89.1%) | 5 (10.9%) | 0 (0.0%) | 41 (89.1%) | 5 (10.9%) | 0 (0.0%) | 41 (89.1%) | 5 (10.9%) | 0 (0.0%) | 0 (0.0%) | 46 |
| 73 | Discussion | 17 | 12 (26.1%) | 34 (73.9%) | 0 (0.0%) | 17 (37.0%) | 29 (63.0%) | 0 (0.0%) | 15 (32.6%) | 31 (67.4%) | 0 (0.0%) | 5 (10.9%) | 46 |
| 74 | Discussion | 17 | 8 (17.4%) | 38 (82.6%) | 0 (0.0%) | 10 (21.7%) | 36 (78.3%) | 0 (0.0%) | 10 (21.7%) | 36 (78.3%) | 0 (0.0%) | 2 (4.3%) | 46 |
| 75 | Other Information | 18 | 40 (87.0%) | 6 (13.0%) | 0 (0.0%) | 45 (97.8%) | 1 (2.2%) | 0 (0.0%) | 39 (84.8%) | 7 (15.2%) | 0 (0.0%) | 7 (15.2%) | 46 |
| 76 | Other Information | 18 | 7 (15.2%) | 38 (82.6%) | 1 (2.2%) | 7 (15.2%) | 34 (73.9%) | 5 (10.9%) | 7 (15.2%) | 39 (84.8%) | 0 (0.0%) | 5 (10.9%) | 46 |
| 77 | Other Information | 18 | 3 (6.5%) | 42 (91.3%) | 1 (2.2%) | 3 (6.5%) | 41 (89.1%) | 2 (4.3%) | 4 (8.7%) | 42 (91.3%) | 0 (0.0%) | 3 (6.5%) | 46 |
| 78 | Other Information | 19 | 44 (95.7%) | 2 (4.3%) | 0 (0.0%) | 45 (97.8%) | 1 (2.2%) | 0 (0.0%) | 46 (100.0%) | 0 (0.0%) | 0 (0.0%) | 3 (6.5%) | 46 |
| 79 | Other Information | 19 | 3 (6.5%) | 43 (93.5%) | 0 (0.0%) | 3 (6.5%) | 43 (93.5%) | 0 (0.0%) | 3 (6.5%) | 43 (93.5%) | 0 (0.0%) | 2 (4.3%) | 46 |
| 80 | Other Information | 20 | 38 (82.6%) | 8 (17.4%) | 0 (0.0%) | 45 (97.8%) | 1 (2.2%) | 0 (0.0%) | 39 (84.8%) | 7 (15.2%) | 0 (0.0%) | 7 (15.2%) | 46 |

2 NR: not reported, NA: not applicable.

Table S1-5: Model agreement with human consensus by component

| Index | Section | Item | Agreement, % |
|---|---|---|---|
| 1 | Title and Abstract | 1 | 100.0 |
| 2 | Introduction | 2 | 100.0 |
| 3 | Introduction | 3 | 91.3 |
| 4 | Methods | 4 | 100.0 |
| 5 | Methods | 4 | 100.0 |
| 6 | Methods | 4 | 100.0 |
| 7 | Methods | 4 | 97.8 |
| 8 | Methods | 4 | 93.5 |
| 9 | Methods | 4a | 76.1 |
| 10 | Methods | 4a | 91.3 |
| 11 | Methods | 4a | 80.4 |
| 12 | Methods | 4a | 95.7 |
| 13 | Methods | 4a | 78.3 |
| 14 | Methods | 4a | 60.9 |
| 15 | Methods | 4a | 84.8 |
| 16 | Methods | 4b | 93.5 |
| 17 | Methods | 4b | 91.3 |
| 18 | Methods | 4b | 91.3 |
| 19 | Methods | 4c | 69.6 |
| 20 | Methods | 4c | 71.7 |
| 21 | Methods | 4c | 100.0 |
| 22 | Methods | 4d | 82.6 |
| 23 | Methods | 4d | 91.3 |
| 24 | Methods | 4d | 84.8 |
| 25 | Methods | 4e | 80.4 |
| 26 | Methods | 5 | 82.6 |
| 27 | Methods | 5 | 87.0 |
| 28 | Methods | 5 | 89.1 |
| 29 | Methods | 5 | 89.1 |
| 30 | Methods | 6a | 97.8 |
| 31 | Methods | 6b | 71.7 |
| 32 | Methods | 6c | 100.0 |
| 33 | Methods | 6c | 97.8 |
| 34 | Methods | 6c | 100.0 |
| 35 | Methods | 6d | 91.3 |
| 36 | Methods | 6e | 95.7 |
| 37 | Methods | 7 | 91.3 |
| 38 | Methods | 7 | 87.0 |
| 39 | Methods | 7 | 97.8 |
| 40 | Methods | 7 | 65.2 |
| 41 | Methods | 8 | 100.0 |
| 42 | Methods | 9a | 76.1 |
| 43 | Methods | 9b | 97.8 |
| 44 | Results | 10a | 76.1 |
| 45 | Results | 10b | 82.6 |
| 46 | Results | 10c | 71.7 |
| 47 | Results | 10d | 60.9 |
| 48 | Results | 10d | 97.8 |
| 49 | Results | 11a | 78.3 |
| 50 | Results | 11b | 97.8 |
| 51 | Results | 11c | 95.7 |
| 52 | Results | 11d | 91.3 |
| 53 | Results | 12a | 93.5 |
| 54 | Results | 12a | 82.6 |
| 55 | Results | 12a | 95.7 |
| 56 | Results | 12a | 63.0 |
| 57 | Results | 12b | 89.1 |
| 58 | Results | 13a | 100.0 |

| Index | Section | Item | Agreement, % |
|---|---|---|---|
| 59 | Results | 13b | 100.0 |
| 60 | Results | 13c | 73.9 |
| 61 | Results | 13d | 65.2 |
| 62 | Results | 13e | 76.1 |
| 63 | Discussion | 14 | 100.0 |
| 64 | Discussion | 15 | 93.5 |
| 65 | Discussion | 15 | 100.0 |
| 66 | Discussion | 15 | 82.6 |
| 67 | Discussion | 16a | 100.0 |
| 68 | Discussion | 16b | 100.0 |
| 69 | Discussion | 16b | 91.3 |
| 70 | Discussion | 16b | 91.3 |
| 71 | Discussion | 16c | 100.0 |
| 72 | Discussion | 17 | 97.8 |
| 73 | Discussion | 17 | 89.1 |
| 74 | Discussion | 17 | 84.8 |
| 75 | Other Information | 18 | 93.5 |
| 76 | Other Information | 18 | 100.0 |
| 77 | Other Information | 18 | 97.8 |
| 78 | Other Information | 19 | 93.5 |
| 79 | Other Information | 19 | 97.8 |
| 80 | Other Information | 20 | 89.1 |

Table S1-6: STROBE-MR reporting by component

| Index | Section | Item | Reported, n(%) | Not Reported, n(%) | Not Applicable, n(%) |
|---|---|---|---|---|---|
| 1 | Title and Abstract | 1 | 12407 (97.7%) | 187 (1.5%) | 105 (0.8%) |
| 2 | Introduction | 2 | 12318 (97.0%) | 336 (2.6%) | 45 (0.4%) |
| 3 | Introduction | 3 | 11930 (93.9%) | 716 (5.6%) | 53 (0.4%) |
| 4 | Methods | 4 | 12543 (98.8%) | 133 (1.0%) | 23 (0.2%) |
| 5 | Methods | 4 | 12530 (98.7%) | 146 (1.1%) | 23 (0.2%) |
| 6 | Methods | 4 | 12532 (98.7%) | 124 (1.0%) | 43 (0.3%) |
| 7 | Methods | 4 | 12564 (98.9%) | 89 (0.7%) | 46 (0.4%) |
| 8 | Methods | 4 | 7016 (55.2%) | 5644 (44.4%) | 39 (0.3%) |
| 9 | Methods | 4a | 7917 (62.3%) | 4749 (37.4%) | 33 (0.3%) |
| 10 | Methods | 4a | 10474 (82.5%) | 2189 (17.2%) | 36 (0.3%) |
| 11 | Methods | 4a | 7823 (61.6%) | 4824 (38.0%) | 52 (0.4%) |
| 12 | Methods | 4a | 1860 (14.6%) | 10790 (85.0%) | 49 (0.4%) |
| 13 | Methods | 4a | 9393 (74.0%) | 3264 (25.7%) | 42 (0.3%) |
| 14 | Methods | 4a | 1555 (12.2%) | 3326 (26.2%) | 7818 (61.6%) |
| 15 | Methods | 4a | 7210 (56.8%) | 5460 (43.0%) | 29 (0.2%) |
| 16 | Methods | 4b | 4121 (32.5%) | 8541 (67.3%) | 37 (0.3%) |
| 17 | Methods | 4b | 11422 (89.9%) | 1256 (9.9%) | 21 (0.2%) |
| 18 | Methods | 4b | 1617 (12.7%) | 11049 (87.0%) | 33 (0.3%) |
| 19 | Methods | 4c | 5805 (45.7%) | 6803 (53.6%) | 91 (0.7%) |
| 20 | Methods | 4c | 8160 (64.3%) | 4434 (34.9%) | 105 (0.8%) |
| 21 | Methods | 4c | 11956 (94.1%) | 653 (5.1%) | 90 (0.7%) |
| 22 | Methods | 4d | 7461 (58.8%) | 5187 (40.8%) | 51 (0.4%) |
| 23 | Methods | 4d | 6699 (52.8%) | 5951 (46.9%) | 49 (0.4%) |
| 24 | Methods | 4d | 5505 (43.3%) | 7106 (56.0%) | 88 (0.7%) |
| 25 | Methods | 4e | 9537 (75.1%) | 3064 (24.1%) | 98 (0.8%) |
| 26 | Methods | 5 | 9918 (78.1%) | 2660 (20.9%) | 121 (1.0%) |
| 27 | Methods | 5 | 9279 (73.1%) | 3299 (26.0%) | 121 (1.0%) |
| 28 | Methods | 5 | 9394 (74.0%) | 3184 (25.1%) | 121 (1.0%) |
| 29 | Methods | 5 | 2008 (15.8%) | 10548 (83.1%) | 143 (1.1%) |
| 30 | Methods | 6a | 5620 (44.3%) | 7045 (55.5%) | 34 (0.3%) |
| 31 | Methods | 6b | 10289 (81.0%) | 2311 (18.2%) | 99 (0.8%) |
| 32 | Methods | 6c | 12163 (95.8%) | 391 (3.1%) | 145 (1.1%) |

| Index | Section | Item | Reported, n(%) | Not Reported, n(%) | Not Applicable, n(%) |
|---|---|---|---|---|---|
| 33 | Methods | 6c | 4622 (36.4%) | 8031 (63.2%) | 46 (0.4%) |
| 34 | Methods | 6c | 454 (3.6%) | 11309 (89.1%) | 936 (7.4%) |
| 35 | Methods | 6d | 2023 (15.9%) | 10620 (83.6%) | 56 (0.4%) |
| 36 | Methods | 6e | 6048 (47.6%) | 6545 (51.5%) | 106 (0.8%) |
| 37 | Methods | 7 | 11040 (86.9%) | 1540 (12.1%) | 119 (0.9%) |
| 38 | Methods | 7 | 7950 (62.6%) | 4630 (36.5%) | 119 (0.9%) |
| 39 | Methods | 7 | 11736 (92.4%) | 844 (6.6%) | 119 (0.9%) |
| 40 | Methods | 7 | 10182 (80.2%) | 2371 (18.7%) | 146 (1.1%) |
| 41 | Methods | 8 | 12271 (96.6%) | 391 (3.1%) | 37 (0.3%) |
| 42 | Methods | 9a | 7509 (59.1%) | 5172 (40.7%) | 18 (0.1%) |
| 43 | Methods | 9b | 304 (2.4%) | 12377 (97.5%) | 18 (0.1%) |
| 44 | Results | 10a | 5997 (47.2%) | 6666 (52.5%) | 36 (0.3%) |
| 45 | Results | 10b | 4615 (36.3%) | 8046 (63.4%) | 38 (0.3%) |
| 46 | Results | 10c | 1215 (9.6%) | 8104 (63.8%) | 3380 (26.6%) |
| 47 | Results | 10d | 4169 (32.8%) | 7637 (60.1%) | 893 (7.0%) |
| 48 | Results | 10d | 2083 (16.4%) | 9737 (76.7%) | 879 (6.9%) |
| 49 | Results | 11a | 8403 (66.2%) | 4186 (33.0%) | 110 (0.9%) |
| 50 | Results | 11b | 11607 (91.4%) | 938 (7.4%) | 154 (1.2%) |
| 51 | Results | 11c | 269 (2.1%) | 11738 (92.4%) | 692 (5.4%) |
| 52 | Results | 11d | 11626 (91.6%) | 1021 (8.0%) | 52 (0.4%) |
| 53 | Results | 12a | 9093 (71.6%) | 3480 (27.4%) | 126 (1.0%) |
| 54 | Results | 12a | 7080 (55.8%) | 5493 (43.3%) | 126 (1.0%) |
| 55 | Results | 12a | 11250 (88.6%) | 1323 (10.4%) | 126 (1.0%) |
| 56 | Results | 12a | 6963 (54.8%) | 5440 (42.8%) | 296 (2.3%) |
| 57 | Results | 12b | 9670 (76.1%) | 2785 (21.9%) | 244 (1.9%) |
| 58 | Results | 13a | 12052 (94.9%) | 585 (4.6%) | 62 (0.5%) |
| 59 | Results | 13b | 11848 (93.3%) | 795 (6.3%) | 56 (0.4%) |
| 60 | Results | 13c | 5903 (46.5%) | 4888 (38.5%) | 1908 (15.0%) |
| 61 | Results | 13d | 8303 (65.4%) | 4233 (33.3%) | 163 (1.3%) |
| 62 | Results | 13e | 7919 (62.4%) | 4473 (35.2%) | 307 (2.4%) |
| 63 | Discussion | 14 | 12587 (99.1%) | 88 (0.7%) | 24 (0.2%) |
| 64 | Discussion | 15 | 9251 (72.8%) | 3344 (26.3%) | 104 (0.8%) |
| 65 | Discussion | 15 | 11969 (94.3%) | 712 (5.6%) | 18 (0.1%) |
| 66 | Discussion | 15 | 9686 (76.3%) | 2995 (23.6%) | 18 (0.1%) |
| 67 | Discussion | 16a | 12416 (97.8%) | 263 (2.1%) | 20 (0.2%) |
| 68 | Discussion | 16b | 12387 (97.5%) | 259 (2.0%) | 53 (0.4%) |
| 69 | Discussion | 16b | 1260 (9.9%) | 11320 (89.1%) | 119 (0.9%) |
| 70 | Discussion | 16b | 10754 (84.7%) | 1839 (14.5%) | 106 (0.8%) |
| 71 | Discussion | 16c | 12323 (97.0%) | 344 (2.7%) | 32 (0.3%) |
| 72 | Discussion | 17 | 10872 (85.6%) | 1805 (14.2%) | 22 (0.2%) |
| 73 | Discussion | 17 | 3314 (26.1%) | 9329 (73.5%) | 56 (0.4%) |
| 74 | Discussion | 17 | 2534 (20.0%) | 10108 (79.6%) | 57 (0.4%) |
| 75 | Other Information | 18 | 10586 (83.4%) | 2101 (16.5%) | 12 (0.1%) |
| 76 | Other Information | 18 | 3087 (24.3%) | 9531 (75.1%) | 81 (0.6%) |
| 77 | Other Information | 18 | 1802 (14.2%) | 10791 (85.0%) | 106 (0.8%) |
| 78 | Other Information | 19 | 11719 (92.3%) | 965 (7.6%) | 15 (0.1%) |
| 79 | Other Information | 19 | 1541 (12.1%) | 11145 (87.8%) | 13 (0.1%) |
| 80 | Other Information | 20 | 10689 (84.2%) | 1998 (15.7%) | 12 (0.1%) |

Table S1-7: Patterns of agreement between model ratings and human consensus

| Pattern | | | |
|---|---|---|---|
| Reviewers agreed? | Model matched consensus? | Example | Interpretation |
| Yes | No | Component 20, *Describe quality control of genetic variants*: the model sometimes treated genetic-variant selection as a quality-control procedure. | Clarify the distinction between variant selection and quality control. |
| Yes | No | Component 31, *Describe how genetic variants were handled in the analyses*: in PMC12450768, the model identified selection thresholds and LD pruning but marked the component as not reported because harmonization and weighting details were absent while both reviewers considered the reported information sufficient. | Specify the minimum evidence needed to satisfy this component. |
| Yes | No | Component 61, *Report and compare with estimates from non-MR analyses*: the model sometimes treated comparison with other observational studies as a comparison with non-MR estimates. | Make it clear that MR authors should include a non-MR analysis in their analysis pipeline and compare its results with MR analysis. |
| Yes | No | Components 14, 46, and 60: the model sometimes assigned ”not reported” where reviewers assigned ”not applicable”, or the reverse. | Provide component-specific guidance on when ”not applicable” should be used. |
| Yes | No | Component 80, *All authors should declare all potential conflicts of interest*: in some cases, a declaration found by reviewers was absent from the extracted text supplied to the model. | We need to make sure the model and human reviewers see the same input. |
| No | Yes | Component 46, *Assess heterogeneity across studies contributing to source meta-analyses*: the model distinguished heterogeneity across source studies from heterogeneity across genetic variants in the MR analysis. | The model can match the final human consensus even when reviewers initially disagree. |
| No | Yes | Component 54, *Report results of the assessment of the independence assumption*: the model distinguished tests of horizontal pleiotropy from assessments of the independence assumption. | Examples of assumptions and procedures to test them may help both human reviewers and automated tools. |

### S2 Supplementary figures

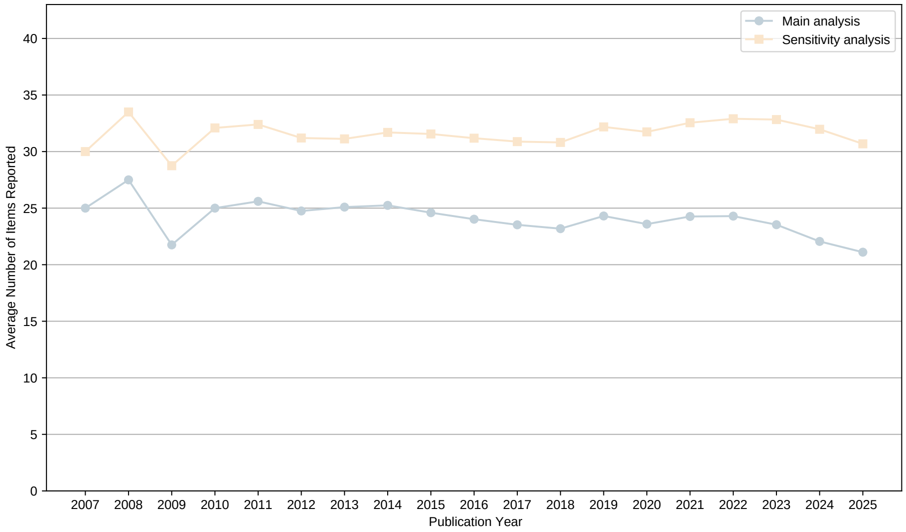


Figure S2-1: Sensitivity analysis: STROBE-MR reporting by publication year

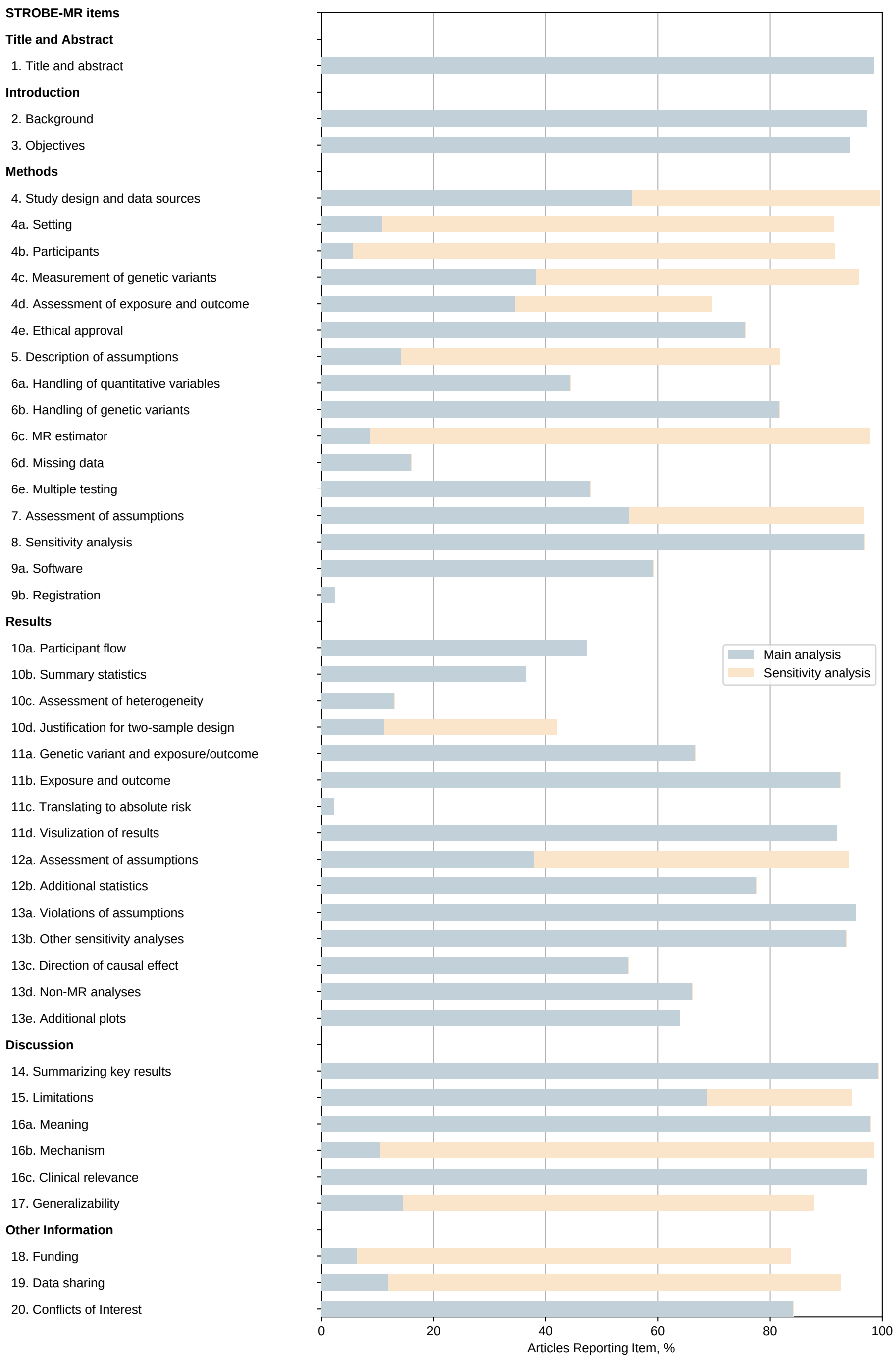


Figure S2-2: Sensitivity analysis: STROBE-MR reporting by item

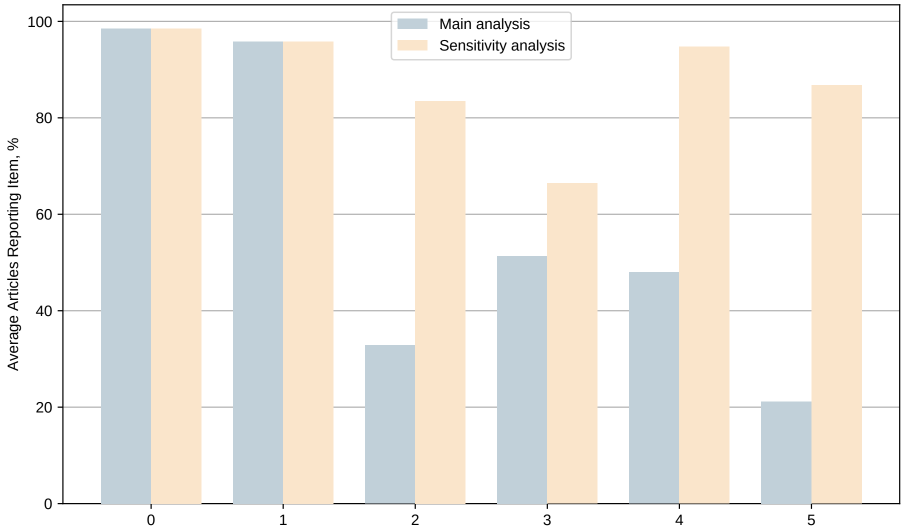


Figure S2-3: Sensitivity analysis: STROBE-MR reporting by section

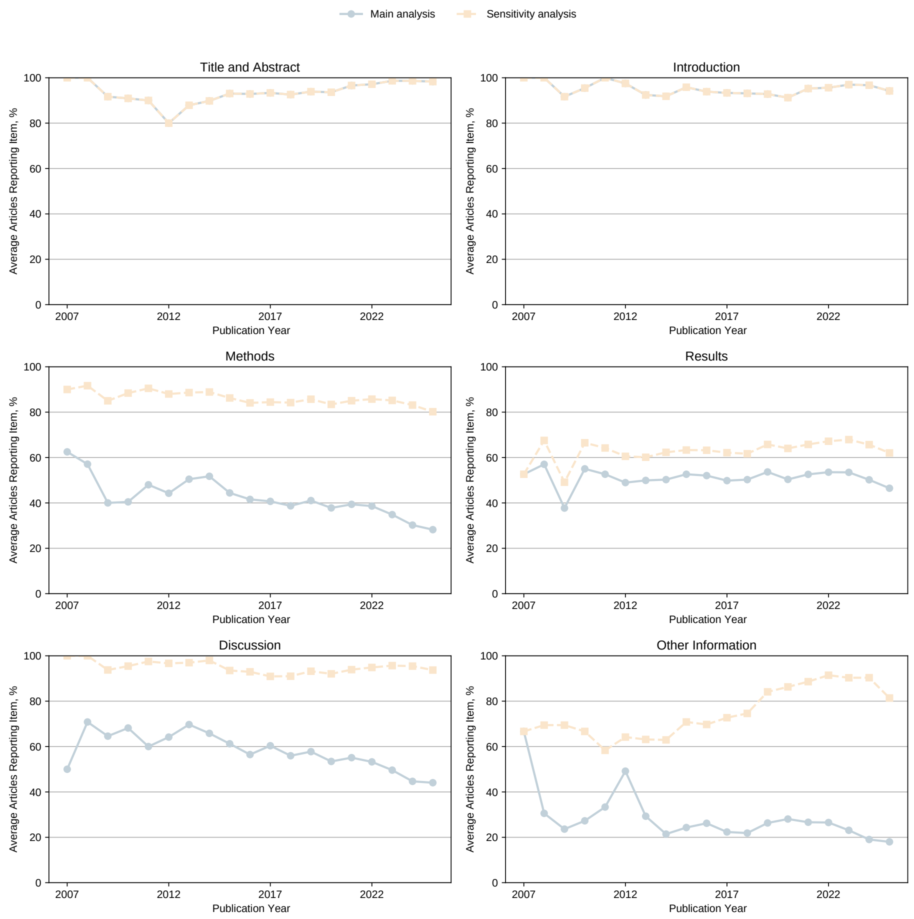


Figure S2-4: Sensitivity analysis: STROBE-MR reporting by year and section

#### S3 Declaration on the use of large language models

In our work, as reported in the main text, LLMs were used as part of the research to assess reporting quality of MR studies.

LLMs have been used as coding agents to assist:

- for research analysis, the implementation of code (Python web crawler and annotation tools) and documentation;
- for the spell and grammar checking of the manuscript.

Coding agents used in our work include GitHub Copilot and Codex. The LLMs used include GPT-5.3-codex and GPT-6-Sol. The LLMs are accessed via API and IDE plugins.

Figures and tables used in the manuscript are generated from analysis code which AI was used in their implementation, but are not directly generated by AI.

Other non-AI / non-LLM tools in proof-reading the manuscript include textidote (`https://github.com/sylvainhalle/textidote`) and typos (`https://github.com/crate-ci/typos`).

The final manuscript is fully drafted by the authors and has been reviewed and edited by all co-authors. Prior to submission we have reviewed ISCB Policy for Acceptable Use of Large Language Models (`https://www.iscb.org/iscb-policy-statements/iscb-policy-for-acceptable-use-of-large-language-models`) and follow its guidelines on the acceptable uses.

#### References
